# Revisiting the Trans-Ancestry Genetic Correlation of Refractive Error

**DOI:** 10.1101/2025.05.08.25327073

**Authors:** Rosie Clark, Xi He, Thu Nga Nguyen, Thanh Huyen Bui, Hannah Noor, Cathy Williams, Louise Terry, Jeremy A. Guggenheim, the UK Biobank Eye and Vision Consortium

## Abstract

**Purpose:** The prevalence of myopia varies significantly across the globe. This may be a consequence of differences in exposure to lifestyle risk factors or differences in genetic susceptibility across ancestry groups. “Trans-ancestry genetic correlation” quantifies the similarity in genetic predisposition to a trait or disease between different populations. We aimed to calculate the trans-ancestry genetic correlation of refractive error across a diverse range of ancestry groups (European, South Asian, East Asian and African) using recently developed approaches.

**Methods:** Two methods were applied: (1) trans-ancestry genetic correlation with unbalanced data resources (TAGC-UDR) and (2) trans-ancestry bivariate genomic-relatedness-based restricted maximum-likelihood (TAB-GREML). TAGC-UDR analyses were carried out for UK Biobank participants of European (n=107,668; n=3,500), East Asian (n=972), South Asian (n=4,303) and African (n=3,877) ancestry. TAB-GREML analyses were carried out for participants of European (n=10,000), South Asian (n=4,303) and African (n=3,877) ancestry.

**Results:** TAGC-UDR analyses suggested the trans-ancestry genetic correlation of refractive error was in the range 0.7–1.0 for the European vs. African, European vs. East Asian and European vs. South Asian ancestry pairs. The TAB-GREML estimates were consistent with the TAGC-UDR findings. Precision of the estimates was limited, reflecting the modest sample sizes of the non-European samples.

**Conclusion:** The findings support existing evidence that genetic susceptibility to refractive error is largely shared between Europeans and East Asians, and further suggest that genetic susceptibility to refractive error is largely shared across Europeans, Africans and South Asians. The results imply that differences in geographical variation in myopia prevalence are mostly driven by lifestyle factors.

## Introduction

Refractive errors occur due to a mismatch between the combined focal power of the cornea and crystalline lens compared to the eye’s axial length^1^. In myopia, the axial length is too long relative to the eye’s optical power, which causes light to focus in front of the plane of the retinal photoreceptors. The converse happens in hyperopia, with light focussing behind the plane of the retinal photoreceptors. The blurry vision produced by myopia and hyperopia can be corrected by wearing glasses or contact lenses, or by refractive surgery. However, myopia and hyperopia are associated with serious comorbidities^2^. For instance, high myopia is a risk factor for glaucoma, retinal detachment and myopic macular degeneration, which are increasingly common causes of visual impairment^3^.

Both genetics and environmental (lifestyle) factors contribute to the development of refractive errors^2^. In certain individuals, early-onset high myopia is caused by a mutation in a single gene. To date, loss of function mutations in approximately 20 different genes have been found to cause non-syndromic high myopia, with *ARR3* and *OPN1LW* being the most frequently reported disease genes^4-7^. More generally, genetic susceptibility to myopia and hyperopia is conferred by inheritance of a very large number of genetic variants; each variant confers a very small increased risk of either myopia or hyperopia, yet in aggregate the combined effect of all of the variants is substantial^8^. More than 450 distinct genetic variants associated with refractive error have been identified by genome-wide association studies^9^.

Currently, the cumulative effect of these genetic variants can explain about 20% of the inter-subject variation in refractive error^10^. Insufficient time spent outdoors during childhood and a high level of education are the most important lifestyle risk factors for myopia that have been discovered^11,12^. Mounting evidence suggests that these lifestyle risk factors trigger the development of myopia in genetically susceptible individuals^2^. However, since the prevalence of myopia in young adults exceeds 80% in some countries, it is likely that even a person with limited genetic susceptibility to myopia will eventually succumb if their exposure to lifestyle risk factors is high enough^2,13^.

A notable feature of the worldwide prevalence of myopia is its geographical variation, exceeding 50% in countries such as China, Taiwan, Hong Kong and Singapore, while being below 20% in Australia and parts of Africa^14,15^. The varied geographical distribution of myopia may result from differences in genetic susceptibility across ancestry groups or differences in the level of exposure to lifestyle risk factors across geographical regions, or a combination of the two. “Trans-ancestry genetic correlation” is a statistical measure used to quantify the similarity in genetic predisposition to a trait or disease across two populations of differing ancestry. A trans-ancestry genetic correlation equal to one implies that, across the genome, genetic variants that confer susceptibility to a disease are shared perfectly between the two ancestral groups. In other words, a trans-ancestry genetic correlation of one implies that genetic variants have the same “effect size” (i.e. the same degree of association with the disease) in the two ancestral populations. By contrast, a trans-ancestry genetic correlation of zero implies unique sources of genetic susceptibility in the two populations. In practice, calculating a measure of trans-ancestry genetic correlation is complicated by differences in allele frequency across ancestry groups (in the extreme case, an allele that confers susceptibility to a disease may be present in one ancestry group but absent from the other).

A range of statistical methods have been developed to estimate trans-ancestry genetic correlations; each method makes different assumptions regarding the effect of allele frequency differences or in its requirement for summary data or individual-level data^16-21^. In past work, the CREAM consortium reported a trans-ancestry genetic correlation for refractive error in European ancestry vs. East Asian ancestry individuals^22^, using an analysis method called *popcorn*^17^. Here, we apply two recently introduced methods for calculating the trans-ancestry genetic correlation of refractive error that rely on different assumptions to *popcorn*. For our primary analysis we apply the Trans-Ancestry Genetic Correlation with Unbalanced Data Resources (TAGC-UDR) method^19^, which is designed for use when the sample size of one of the two ancestral populations is limited. As a sensitivity analysis we apply the trans-ancestry bivariate genomic-relatedness-based restricted maximum-likelihood (TAB-GREML) method, which fits a random effects model similar to the widely-employed bivariate GREML approach^23^, but that takes into account differences in allele frequency between the two ancestry samples rather than assuming a common allele frequency^21,24^. We build on the existing CREAM findings by estimating the trans-ancestry genetic correlation of refractive error for a diverse set of ancestry groups: European, East Asian, South Asian, and African.

## Methods

### Overview of the study methods

We applied two methods of calculating trans-ancestry genetic correlations: TAGC-UDR and TAB-GREML. The TAGC-UDR method was used as our primary analysis because this approach has been reported to perform well when one ancestry sample is much larger than the other^19^. TAB-GREML was used as a sensitivity analysis; this approach requires relatively large sample sizes for both ancestry groups and can be biased if the sample sizes are very highly imbalanced^19^.

#### TAGC-UDR

Unrelated UK Biobank participants with information available for refractive error were classified into one of five ancestry groups: European (n=111,168), East Asian (n=972), South Asian (n=4,303), African (n=3,877), and Other. The European sample was split into a GWAS sample (n=107,668) and an “evaluation sample” (n=3,500). A GWAS for refractive error was performed in the European ancestry GWAS sample (n=107,668), using a subset of HapMap3 genetic variants^25^. The regression coefficients for all variants included in the GWAS were used to derive weights for a polygenic score, which was then applied to participants in the four evaluation samples: European (n=3,500), East Asian (n=972), South Asian (n=4,303) and African (n=3,877). Trans-ancestry “bias factors” ^19^ were computed using linkage disequilibrium (LD) reference samples from each ancestry group combined with an estimate of the single nucleotide polymorphism (SNP)-heritability 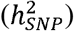 of refractive error in each ancestry group (since SNP-heritability is challenging to calculate in small samples, calculations were performed using a range of plausible values: 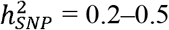). The role of the bias factors is to up-scale the polygenic score in the evaluation sample to account for (i) imprecision in GWAS beta coefficient estimation due to the finite size of the GWAS analysis, and (ii) differences in LD-structure between ancestry groups^19^. Comparison of the true refractive error vs. the up-scaled genetically predicted refractive error for participants in the evaluation samples was used to estimate the trans-ancestry genetic correlation. Standard errors were estimated by bootstrapping.

#### TAB-GREML

analysis is a modification of standard bivariate GREML analysis^23^ that takes into account the differences in allele frequency in two ancestry samples when calculating trans-ancestry genetic correlations^21,24^. Unrelated UK Biobank participants with information available for refractive error were classified into one of five ancestry groups as described in the TAGC-UDR section above. Next, a set of 10,000 European ancestry participants was selected at random from the full European-ancestry sample. TAB-GREML analyses were carried out for (a) n=10,000 European and n=4,303 South Asian ancestry participants, and (b) the n=10,000 European and n=3,877 African ancestry participants. The sample size of East Asian ancestry participants (n=972) was too small to fit a TAB-GREML model, which prevented a European-East Asian trans-ancestry genetic correlation being calculated using the TAB-GREML method.

### Selection of participants from the UK Biobank cohort

Ethical approval for the UK Biobank study was obtained from the NHS Research Ethics Committee (Reference: 11/NW/0382). Approximately 500,000 participants aged 40-70 years-old were recruited from across the United Kingdom, during the period 2006 and 2010, via one of twenty-two assessment centres^26^. Approximately one quarter of the UK Biobank cohort had their refractive error measured (Tomey RC 5000 autorefractor, Tomey Corp., Nagoya, Japan). Blood samples were collected and genotyped at ∼800,000 markers using either the UK BiLEVE Axiom array or the UK Biobank Axiom array. Imputation to a joint Haplotype Reference Consortium–UK 10,000 Genomes Project reference panel was performed by Bycroft et al.^27^. Participants were included in the current analysis if they had information available for their refractive error, were seen at an assessment centre at which more than n=100 participants had refractive error assessed, and had a genetic heterozygosity within 10 standard deviations of the mean for the full sample. Ancestry was classified using a loose definition based solely on genetic principal components 1 and 2 (PC1 and PC2) with the aim of maximizing the sample size of each ancestry group. Specifically, Europeans were defined as individuals with PC1 and PC2 within 10 standard deviations of the mean for participants who self-reported their ethnicity as “White British”. East Asians were defined as individuals with PC1 and PC2 within 5 standard deviations of the mean for participants who self-reported their ethnicity as “Chinese”. South Asians were defined as individuals with PC1 and PC2 within 1.25 standard deviations of the mean for participants who self-reported their ethnicity as “Asian”. Africans were defined as individuals with PC1 and PC2 within 2 standard deviations of the mean for participants who self-reported their ethnicity as “Black”. Figure 1 illustrates the distribution of PC1 and PC2 in each ancestry group. Within each of the African, East Asian and South Asian groups, a maximal set of unrelated participants was selected using a custom R script that applied the R package *igraph* function to the kinship data reported by Bycroft et al.^27^. This resulted in samples of unrelated African (n=3,877), East Asian (n=972) and South Asian (n=4,303) participants, which we refer to as “evaluation samples”. For the European ancestry sample, a set of n=3,500 unrelated participants was first selected at random as a European “evaluation sample”. The remaining n=107,668 European ancestry participants were used as a GWAS sample. From within the African, East Asian, European, and South Asian evaluation samples, a set of approximately 1000 unrelated participants was selected at random to serve as an LD reference sample for that ancestry group.

**Figure 1.**
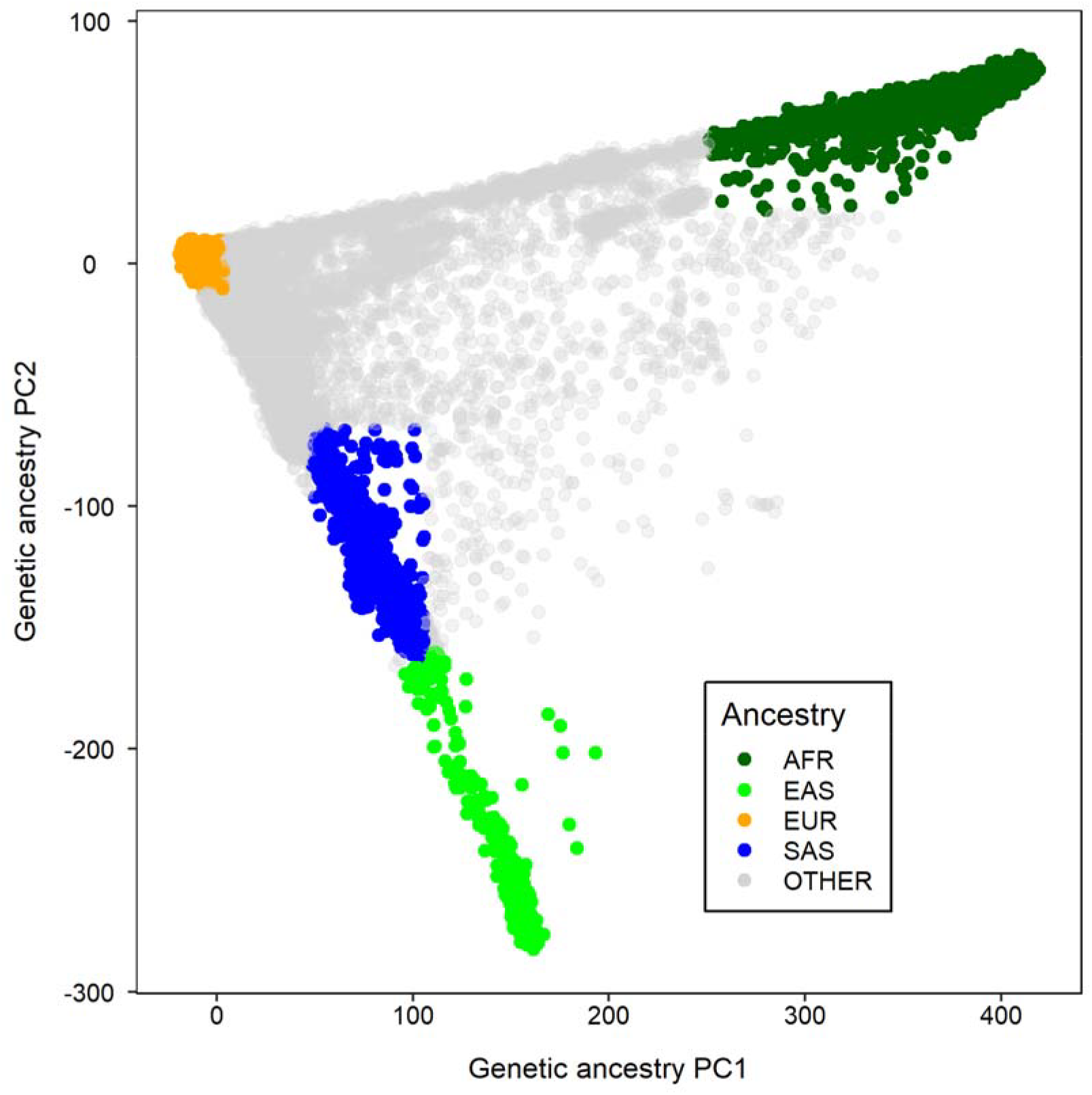
Genetic ancestry classification based on genetic principal components 1 and 2 (PC1 and PC2). In comparison to EUR participants, wider distributions of PC1 and PC2 were used to define AFR, EAS and SAS participants in order to maximise the available sample sizes. Abbreviations: AFR = African, EAS = East Asian, EUR = European, SAS = South Asian.

### Selection of genetic variants

The set of n= 990,761 HapMap3 variants reported by Zhao et al.^19^ (https://github.com/FSSKM/TAGC_review/blob/main/TAGC_data/hm3_snp.id.list) was filtered to remove: (i) variants with a Hardy Weinberg equilibrium test *P* < 1e-04 in any of the four LD reference ancestry samples, (ii) variants with a minor allele frequency (MAF) < 1% in any of the four LD reference ancestry samples, and (iii) variants with a genotyping rate < 90% in any of the four LD reference ancestry samples. This resulted in a final set of n=642,807 HapMap3 variants.

### Genome-wide association study for refractive error

A GWAS for autorefractor-measured spherical equivalent refractive error (in Diopters) averaged between the two eyes was performed using BOLT-LMM v2.4.1 ^28^ for the n=107,668 European ancestry GWAS sample. Age, age^2, sex, genotyping array, assessment center, and genetic ancestry PC1-10 were included as covariates. The n=642,807 HapMap3 variants were analyzed.

### Trans-ancestry genetic correlation analysis with TAGC-UDR

Using the terminology from Zhao et al.^19^, the 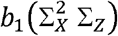and 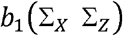 LD block matrix moment terms were calculated for each of the 253 LD blocks identified by Zhao et al.^19^ in each ancestry pair (African-European, East Asian-European, South Asian-European, European-European) with the *TAGC_LD_block_moment* and *WAGC_LD_block_moment* functions from the R package *TAGC* (https://github.com/FSSKM/TAGC_review). These LD moments were then pooled across the genome for each ancestry pair and used to calculate trans-ancestry bias factors for each ancestry pair^19^.

A polygenic score for refractive error was calculated using PLINK v1.90b6 ^29^ for participants in each of the four evaluation samples, using the SNP weights from the GWAS for autorefractor-measured spherical equivalent refractive error described above. The polygenic score and the autorefractor-measured refractive error were both standardized (transformed to have mean=0, standard deviation=1) within each evaluation sample. Then, standardized autorefractor-measured refractive error was regressed on the standardized polygenic score, and a set of standardized covariates (age, age^2, sex, and PC1-PC10) using 500 bootstrap replicates. The regression coefficient and its bootstrap standard error were both multiplied by the ancestry-specific bias factor to yield an estimate of the trans-ancestry genetic correlation^19^.

### Sensitivity analysis: Trans-ancestry genetic correlation analysis with TAB-GREML

Sets of unrelated UK Biobank participants of African (n=3,877) and South Asian (n=4,303) ancestry were selected as described in the *Sample selection* section above. From the full set of European-ancestry participants (n=111,168) described in the *Sample selection* section above, n=10,000 unrelated participants were chosen at random. A genetic relationship matrix (GRM) was constructed for the combined African + European sample (n=13,877) and for the combined South Asian + European sample (n=14,303), using the *-rtmx2* command of the MTG2 software^21^. TAB-GREML analysis was performed using the *-mod 2* command of MTG2, with autorefractor-measured refractive error as the phenotype and age, age^2, sex, and PC1-PC10 as covariates.

## Results

Demographic characteristics of the European-ancestry GWAS sample and the four multi-ancestry evaluation samples are presented in Table 1. All participants were adults aged 40-70 years. The African, East Asian and South Asian evaluation samples were all younger, on average, than the European evaluation sample (Mann-Whitney U-test; all *P* < 0.001). The median level of refractive error in the East Asian evaluation sample was negative, while it was positive in the other samples. Indeed, refractive error was significantly more myopic in the East Asian sample compared to the other three evaluation samples (Mann-Whitney U-test; all *P* < 0.001). In accordance with the more myopic refractive error in the East Asian sample, this ancestry group’s self-reported age of first wearing glasses/contact lenses was younger than that of the other three ancestry samples (Mann-Whitney U-test; all *P* < 0.001). These ancestry-specific refractive error differences have been observed previously in UK Biobank participants^30^.

**Table 1.** Demographic characteristics of the GWAS sample and the multi-ancestry evaluation samples.

| **Evaluation sample** | **European**<br>**GWAS sample** | **African**<br>**Evaluation sample** | **East Asian**<br>**Evaluation sample** | **European**<br>**Evaluation sample** | **South Asian**<br>**Evaluation sample** |
| --- | --- | --- | --- | --- | --- |
| Sample size | 107,668 | 3,877 | 972 | 3,500 | 4,303 |
| Female<br>percentage | 53.4% | 59.1% | 68.0% | 54.1% | 48.2% |
| Age (years)<br>median (Q1-Q3) | 59.83<br>(52.17 to 64.50) | 51.67<br>(46.08 to 58.58) | 53.83<br>(47.17 to 60.42) | 60.00<br>(52.24 to 64.83) | 54.17<br>(46.71 to 61.33) |
| Age of first spectacles<br>(years)<sup>†</sup><br>median (Q1-Q3) | 37.00<br>(15.00 to 47.00) | 37.50<br>(19.00 to 45.00) | 25.00<br>(14.00 to 45.00) | 38.00<br>(15.00 to 48.00) | 40.00<br>(18.00 to 45.00) |
| Refractive error (D)<br>median (Q1-Q3) | +0.14<br>(-1.23 to 1.14) | +0.04<br>(-0.82 to 0.70) | -0.44<br>(-2.98 to 0.45) | +0.16<br>(-1.18 to 1.14) | +0.02<br>(-1.05 to 0.76) |
| <sup>†</sup>Age of first spectacles had a bi-modal distribution with peaks at around 13 years-old and 50 years-old, reflecting correction for myopia and presbyopic, respectively. |  |  |  |  |  |

Figure 2 shows the trans-ancestry genetic correlation of refractive error across pairs of ancestry groups calculated using the TAGC-UDR method (further details can be found in Supplementary Figure S1). The European vs. European comparison was included as a positive control: an evaluation of participants with the same genetic ancestry would be expected to have a genetic correlation of *r*_*g*_ = 1.0. Estimation of the trans-ancestry genetic correlation requires the “SNP-heritability” of refractive error in each ancestry sample to be specified (SNP-heritability refers to the genetic contribution to a trait that can be explained by a particular set of SNPs^31^). In Figure 2, heritability was assumed to be equal in each pair of ancestry groups being evaluated; this assumption was relaxed for the analyses shown in Supplementary Figure S1. Since a precise SNP-heritability value is challenging to estimate in small samples^23^ and can vary depending on the specific set of SNPs being considered, we calculated the trans-ancestry genetic correlations across a range of plausible heritability values. At each potential SNP-heritability value, the estimated trans ancestry genetic correlation was nominally higher in the European vs. European ancestry pair, followed by the European vs. South Asian ancestry pair, European vs. East Asian ancestry pair and finally the European vs. African ancestry pair (Figure 2). However, the 95% confidence intervals of these pairwise estimates overlapped, suggesting that the trans-ancestry genetic correlation estimates were similar, given the level of precision achieved by the TAGC-UDR analysis method.

**Figure 2.**
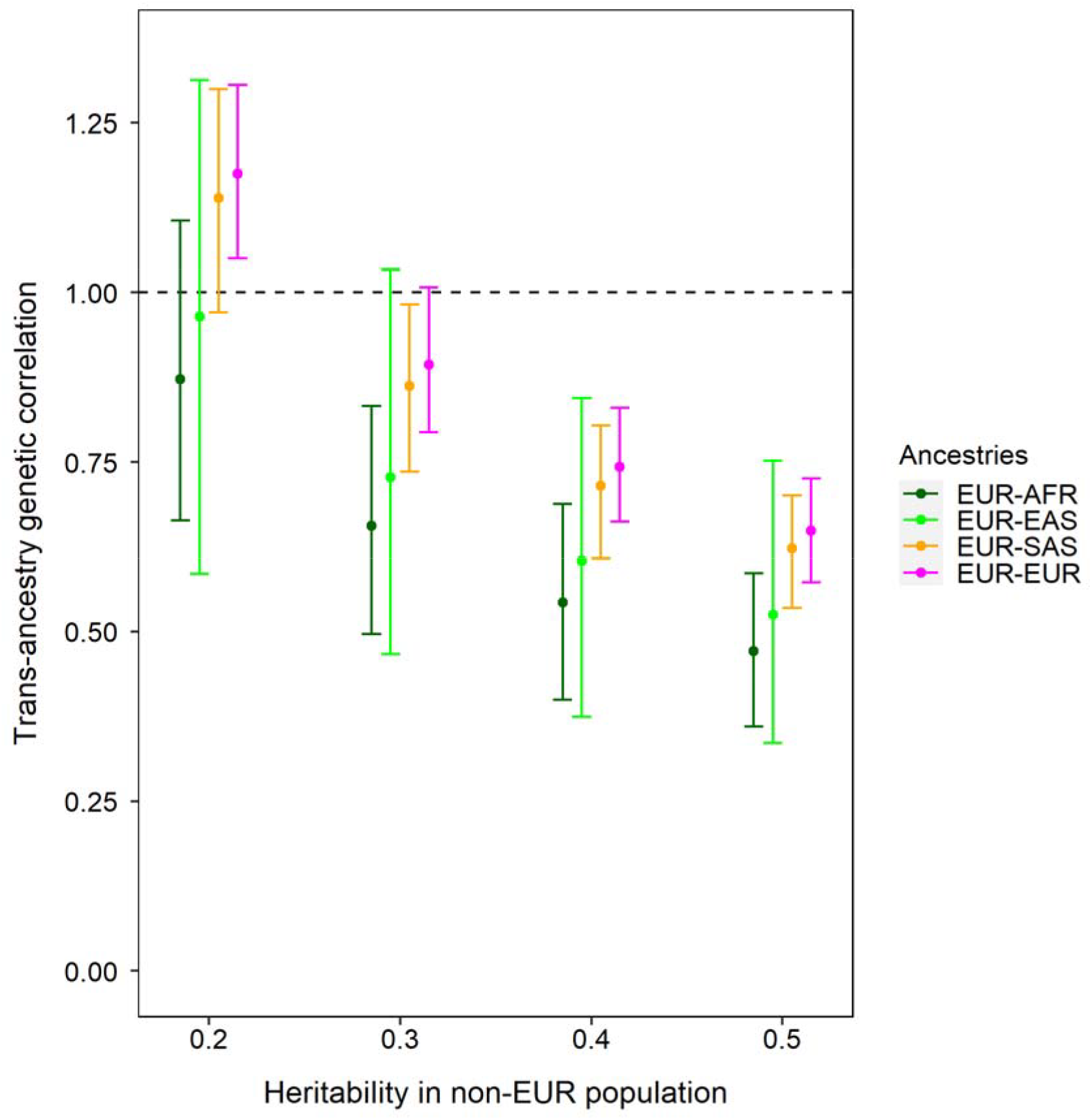
Trans-ancestry genetic correlation estimates. Error bars correspond to 95% confidence intervals. The EUR-EUR comparison was included as a positive control, with the expectation that this would provide a genetic correlation *r*_*g*_ ≈ 1.0. Abbreviations: AFR = African, EAS = East Asian, EUR = European, SAS = South Asian.

When the SNP-heritability of refractive error was assumed to be 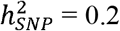, the trans-ancestry genetic correlation estimates were close to *r*_*g*_ ≈ 1.0 for all ancestry pairs. When the SNP-heritability of refractive error was assumed to be higher, the trans-ancestry genetic correlation estimates fell; at the highest SNP heritability level of 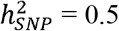, the trans-ancestry genetic correlation was estimated to be *r*_*g*_ ≈ 0.6 for all ancestry pairs. In Figure 2, a genetic correlation of *r*_*g*_ ≈ 1.0 for the European vs. European ancestry pair – matching that expected for this positive control comparison – was observed for SNP-heritability values 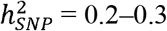. For this (optimal) SNP-heritability level, the trans-ancestry genetic correlation of refractive error was in the range *r*_*g*_ = 0.8–1.0 for the European vs. African, European vs. East Asian and European vs. South Asian ancestry pairs (Table 2).

**Table 2.**
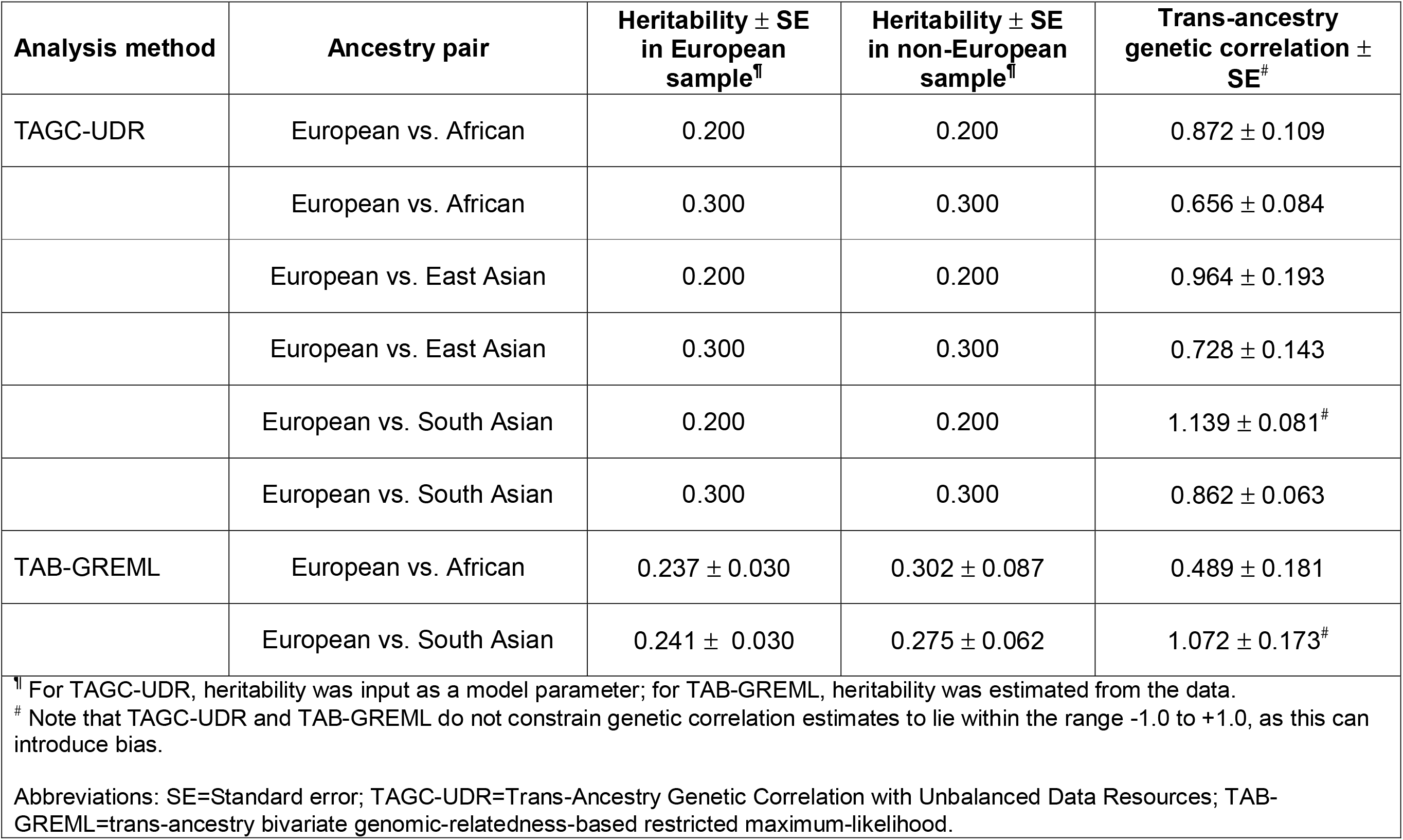
Trans-ancestry genetic correlation estimates for refractive error.

| **Analysis method** | **Ancestry pair** | **Heritability ± SE**<br>**in European**<br>**sample<sup>¶</sup>** | **Heritability ± SE**<br>**in non-European**<br>**sample<sup>¶</sup>** | **Trans-ancestry**<br>**genetic correlation ±**<br>**SE<sup>#</sup>** |
| --- | --- | --- | --- | --- |
| TAGC-UDR | European vs. African | 0.200 | 0.200 | 0.872 ± 0.109 |
| | European vs. African | 0.300 | 0.300 | 0.656 ± 0.084 |
| | European vs. East Asian | 0.200 | 0.200 | 0.964 ± 0.193 |
| | European vs. East Asian | 0.300 | 0.300 | 0.728 ± 0.143 |
| | European vs. South Asian | 0.200 | 0.200 | 1.139 ± 0.081<sup>#</sup> |
| | European vs. South Asian | 0.300 | 0.300 | 0.862 ± 0.063 |
| TAB-GREML | European vs. African | 0.237 ± 0.030 | 0.302 ± 0.087 | 0.489 ± 0.181 |
| | European vs. South Asian | 0.241 ± 0.030 | 0.275 ± 0.062 | 1.072 ± 0.173<sup>#</sup> |
| <sup>¶</sup> For TAGC-UDR, heritability was input as a model parameter; for TAB-GREML, heritability was estimated from the data.<br><sup>#</sup> Note that TAGC-UDR and TAB-GREML do not constrain genetic correlation estimates to lie within the range -1.0 to +1.0, as this can introduce bias.<br>Abbreviations: SE=Standard error; TAGC-UDR=Trans-Ancestry Genetic Correlation with Unbalanced Data Resources; TAB-GREML=trans-ancestry bivariate genomic-relatedness-based restricted maximum-likelihood. |  |  |  |  |

As a sensitivity analysis, the trans-ancestry genetic correlation for the European vs. African and European vs. South Asian ancestry pairs was calculated using an alternative method, TAB-GREML. As shown in Table 2, the TAB-GREML estimates were *r*_*g*_ = 0.49 (standard error [SE] = 0.18) for the European vs. African ancestry pair and *r*_*g*_ = 1.07 (SE = 0.17) for the European vs. South Asian ancestry pair. The high trans-ancestry genetic correlation for the European vs. South Asian ancestry pair supported the result obtained using TAGC-UDR. The TAB-GREML trans-ancestry genetic correlation for the European vs. African ancestry pair was lower than that estimated with TAGC-UDR (TAB-GREML: *r*_*g*_ = 0.49; TAGC-UDR: *r*_*g*_ = 0.66 to 0.87; Table 2). However, these estimates all had large standard errors, suggesting the differences could be due to lack of precision.

## Discussion

Our analyses suggested the trans-ancestry genetic correlation of refractive error lay within the range *r*_*g*_ = 0.7–1.0 across a wide range of ancestry groups – Europeans, Africans, East Asians and South Asians – implying that geographic variation in the prevalence of myopia is mainly driven by differences in lifestyle rather than genetics. The results support the high genetic correlation of refractive error found between European and East Asian individuals by the CREAM consortium (*r*_*g*_ = 0.8–0.9), which was calculated using different sets of participants to those studied here and using an alternative method^22^. Our work extends the earlier CREAM findings by estimating the trans-ancestry genetic correlation across a wider range of ancestries; to our knowledge, the current study is the first to reveal that genetic susceptibility to refractive error is largely shared across Europeans, Africans and South Asians.

When performing a within-ancestry genetic correlation analysis – for example, the correlation of SNP effect sizes for the traits refractive error and axial length in European samples^32^ – the allele frequency of the SNPs with phenotypic effects will generally be well-matched for any two samples of European ancestry. By contrast, in a trans-ancestry genetic correlation analysis, many SNPs may be common in one ancestry group but rare in the other. A SNP that has the same effect size but differs in allele frequency across two ancestral populations will have a greater impact on the phenotype in the ancestry group with the higher allele frequency^16^. Brown et al.^17^ coined the terms “trans-ethnic genetic-impact correlation” (*ρ*_*gi*_) and “trans-ethnic genetic-effect correlation” (*ρ*_*ge*_) to distinguish between genetic correlation estimates that do or do not account for ancestry-specific allele frequency differences, respectively. The TAGC-UDR and TAG-GREML methods we employed here produce estimates that are more akin to *ρ*_*ge*_ than *ρ*_*gi*_.

A key advantage of the TAGC-UDR method for estimating trans-ancestry genetic correlations is that a large sample of participants is only required for one ancestry group (usually, this will be a group of European-ancestry participants, due to the availability of biobank-scale resources for Europeans). Thus, the method can be applied when the available sample size of the second ancestry groups is limited to a few hundred or a few thousand participants^19^. A disadvantage of the TAGC-UDR method is that it produces imprecise estimates, which is a consequence of the small sample size of the second ancestry group. Furthermore, although TAGC-UDR estimates differences in LD patterns across ancestry groups genome-wide, this information is summarized as a set of three LD block moment values (one of which corresponds to the number of SNPs included in the analysis). This approach, while shown to be unbiased in simulations^19^, may not capture subtle trans-ancestry differences in LD structure. Following Momin et al.^21^, we restricted the HapMap3 SNPs used in our analysis to those with a MAF of at least 1% in the ancestry groups studied, to avoid the situation where an allele was essentially monomorphic in one ancestry sample. This meant that our genetic correlation was based on a selected sample of genetic variants, whereas ideally a genetic correlation estimate would be calculated using the set of all causal variants in the genome for the trait-of-interest. Furthermore, due to the limited sample size of the evaluation samples, we inferred the heritability of refractive error by observing the genetic correlation of the European vs. European pair, rather than estimating the heritability directly from the data. As expected theoretically^19^, trans-ancestry genetic correlation estimates were lower when the heritability of refractive error was assumed to be higher (Figure 2). This pattern occurs because precision in estimating SNP effect sizes is worse for traits with low heritability, therefore a pre-determined bias-correction will up-scale effect sizes for traits with low heritability more than for traits with high heritability.

The European sample in our analyses had a median age that was 6–8 years older than the other ancestry samples (Table 1). However, this difference in age would be unlikely to have had a major impact on our results, since (i) we included age as a covariate in our analyses, (ii) by adulthood, both the genetic and environmental contribution to refractive error will have become fully manifest, and (iii) refractive error is relatively stable across the 40-70 year age range^33^. Thus, GWAS regression coefficients would be largely stable across the age range of UK Biobank participants. The East Asian sample in our analysis had a median refractive error that was more negative (myopic) than that of the other ancestry samples. However, this would have been unlikely to affect our genetic correlation estimates since heritability and genetic correlation are based on within-sample variations in trait values, i.e. changing the mean value of a trait does not alter the heritability or genetic correlation estimates for the trait.

In summary, genetic correlations for refractive error across European-African, European-East Asian, and European-South Asian ancestry pairs were estimated to lie in the range *r*_*g*_ = 0.7– 1.0. The current result for European vs. East Asian ancestry individuals matched that obtained by the CREAM consortium using an alternative method. The high trans-ancestry genetic correlations suggest that wide variation in the prevalence of myopia across geographic regions is mainly due to differences in exposure to lifestyle risk factors, not genetics. Nevertheless, small sample sizes for the non-European ancestry samples limited the precision of the current genetic correlation estimates, which indicates a need for biobank-scale collection of genotyped individuals with known refractive error across diverse ancestries. A recent consensus statement from the International Myopia Summit^34^ stressed that public health strategies aimed at reducing visual impairment caused by myopia need to have a global perspective. Our findings suggest that public health efforts targeting lifestyle risk factors for myopia should be effective across a wide range of geographical regions.

## Acknowledgements

This research has been conducted using the UK Biobank Resource (application #83325). UK Biobank was established by the Wellcome Trust; the UK Medical Research Council; the Department for Health (London, UK); Scottish Government (Edinburgh, UK); and the Northwest Regional Development Agency (Warrington, UK). It also received funding from the Welsh Assembly Government (Cardiff, UK); the British Heart Foundation; and Diabetes UK. Collection of eye and vision data was supported by The Department for Health through an award made by the NIHR to the Biomedical Research Centre at Moorfields Eye Hospital NHS Foundation Trust, and UCL Institute of Ophthalmology, London, United Kingdom (grant no. BRC2_009). Additional support was provided by The Special Trustees of Moorfields Eye Hospital, London, United Kingdom (grant no. ST 12 09). Data analysis was carried out using the HAWK computing cluster, maintained by Supercomputing Wales and Cardiff University ARCCA.

## Data Availability

UK Biobank data are available via application at https://www.ukbiobank.ac.uk.

## Author Contributions

Conceptualization: JAG, CW, LT. Formal analysis: JAG. Interpretation: All authors. Writing - original draft: JAG. Writing - review & editing: All authors.

---

**Figure S1.**
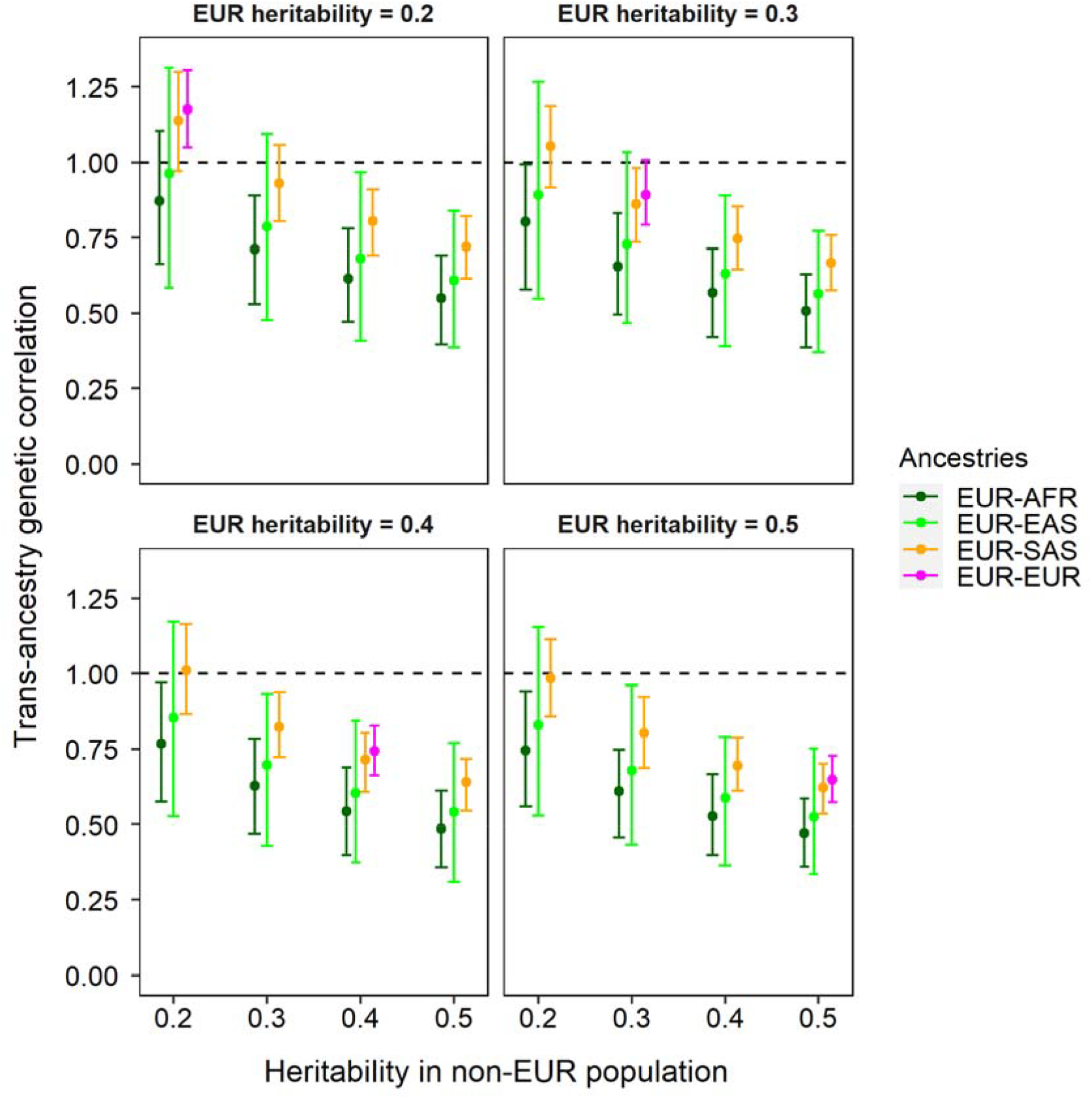
Trans-ancestry genetic correlation estimates. Error bars correspond to 95% confidence intervals. The EUR-EUR comparison was included as a positive control, with the expectation that this would provide a genetic correlation *r*_*g*_ ≈ 1.0. Abbreviations: AFR = African, EAS = East Asian, EUR = European, SAS = South Asian.

## References

1. Baird PN, Saw S-M, Lanca C, et al. Myopia. Nat Rev Dis Primers. 2020;6:99.

2. National Academies of Sciences Engineering and Medicine. Myopia: Causes, Prevention, and Treatment of an Increasingly Common Disease. Washington, DC, USA: The National Academies Press; 2024.

3. Fang Y, Yokoi T, Nagaoka N, et al. Progression of Myopic Maculopathy during 18-Year Follow-up. Ophthalmol. 2018;125:863–877.

4. Gardner JC, Liew G, Quan YH, et al. Three different cone opsin gene array mutational mechanisms; genotype-phenotype correlation and functional investigation of cone opsin variants. Human Mutation. 2014;35:1354–1362.

5. Xiao X, Li S, Jia X, Guo X, Zhang Q. X-linked heterozygous mutations in ARR3 cause female-limited early onset high myopia. Mol Vis. 2016;22:1257–1266.

6. Wang Y, Xiao X, Li X, et al. Genetic and clinical landscape of ARR3-associated MYP26: the most common cause of Mendelian early-onset high myopia with a unique inheritance. Br J Ophthalmol. 2022;doi: 10.1136/bjo-2022-321511.

7. Wang Y, Sun W, Xiao X, et al. Unique Haplotypes in OPN1LW as a Common Cause of High Myopia With or Without Protanopia: A Potential Window Into Myopic Mechanism. Invest Ophthalmol Vis Sci. 2023;64:29.

8. Tideman JWL, Pärssinen O, Haarman AEG, et al. Evaluation of Shared Genetic Susceptibility to High and Low Myopia and Hyperopia. JAMA Ophthalmology. 2021;139:601–609.

9. Hysi PG, Choquet H, Khawaja AP, et al. Meta-analysis of 542,934 subjects of European ancestry identifies new genes and mechanisms predisposing to refractive error and myopia. Nat Genet. 2020;52:401–407.

10. Clark R, Lee SS-Y, Du R, et al. A new polygenic score for refractive error improves detection of children at risk of high myopia but not the prediction of those at risk of myopic macular degeneration. EBioMedicine. 2023;91:104551.

11. Yang Y, Liao H, Zhao L, et al. Green Space Morphology and School Myopia in China. JAMA Ophthalmol. 2024;142:115–122.

12. Morgan IG, Wu P-C, Ostrin LA, et al. IMI Risk Factors for Myopia. Invest Ophthalmol Vis Sci. 2021;62:3.

13. Wojciechowski R. Nature and nurture: the complex genetics of myopia and refractive error. Clinical Genetics. 2011;79:301–320.

14. Attebo K, Ivers RQ, Mitchell P. Refractive errors in an older population: the Blue Mountains Eye Study. Ophthalmol. 1999;106:1066–1072.

15. Wickremasinghe S, Foster PJ, Uranchimeg D, et al. Ocular biometry and refraction in Mongolian adults. Invest Ophthalmol Vis Sci. 2004;45:776–783.

16. Lee SH, Yang J, Goddard ME, Visscher PM, Wray NR. Estimation of pleiotropy between complex diseases using SNP-derived genomic relationships and restricted maximum likelihood. Bioinformatics. 2012;28:2540–2542.

17. Brown Brielin C, Ye Chun J, Price Alkes L, Zaitlen N. Transethnic Genetic-Correlation Estimates from Summary Statistics. American Journal of Human Genetics. 2016;99:76–88.

18. Cai M, Xiao J, Zhang S, et al. A unified framework for cross-population trait prediction by leveraging the genetic correlation of polygenic traits. The American Journal of Human Genetics. 2021;108:632–655.

19. Zhao B, Yang X, Zhu H. Estimating trans-ancestry genetic correlation with unbalanced data resources. J Am Stat Assoc. 2024;119:839–850.

20. Mester R, Hou K, Ding Y, et al. Impact of cross-ancestry genetic architecture on GWASs in admixed populations. The American Journal of Human Genetics. 2023;110:927–939.

21. Momin MM, Shin J, Lee S, et al. A method for an unbiased estimate of cross-ancestry genetic correlation using individual-level data. Nat Commun. 2023;14:722.

22. Tedja MS, Wojciechowski R, Hysi PG, et al. Genome-wide association meta-analysis highlights light-induced signaling as a driver for refractive error. Nat Genet. 2018;50:834–848.

23. Yang JA, Lee SH, Goddard ME, Visscher PM. GCTA: A tool for genome-wide complex trait analysis. American Journal of Human Genetics. 2011;88:76–82.

24. Guo J, Bakshi A, Wang Y, et al. Quantifying genetic heterogeneity between continental populations for human height and body mass index. Scientific Reports. 2021;11:5240.

25. Altshuler DM, Gibbs RA, Peltonen L, et al. Integrating common and rare genetic variation in diverse human populations. Nature. 2010;467:52–58.

26. Sudlow C, Gallacher J, Allen N, et al. UK Biobank: An Open Access Resource for Identifying the Causes of a Wide Range of Complex Diseases of Middle and Old Age. PLoS Med. 2015;12:e1001779.

27. Bycroft C, Freeman C, Petkova D, et al. The UK Biobank resource with deep phenotyping and genomic data. Nature. 2018;562:203–209.

28. Loh P-R, Tucker G, Bulik-Sullivan BK, et al. Efficient Bayesian mixed-model analysis increases association power in large cohorts. Nat Genet. 2015;47:284–290.

29. Chang CC, Chow CC, Tellier LC, et al. Second-generation PLINK: rising to the challenge of larger and richer datasets. GigaScience. 2015;4:7.

30. Cumberland PM, Bao Y, Hysi PG, et al. Frequency and Distribution of Refractive Error in Adult Life: Methodology and Findings of the UK Biobank Study. PLoS ONE. 2015;10:e0139780.

31. Guggenheim JA, St Pourcain B, McMahon G, et al. Assumption-free estimation of the genetic contribution to refractive error across childhood. Mol Vis. 2015;21:621–632.

32. Jiang C, Melles RB, Yin J, et al. A multiethnic genome-wide analysis of 19,420 individuals identifies novel loci associated with axial length and shared genetic influences with refractive error and myopia. Front Genet. 2023;14:1113058.

33. Weale RA. Epidemiology of refractive errors and presbyopia. Survey of ophthalmology. 2003;48:515–543.

34. Furukawa T, Morrow EM, Cepko CL. Crx, a Novel otx-like Homeobox Gene, Shows Photoreceptor-Specific Expression and Regulates Photoreceptor Differentiation. Cell. 1997;91:531–541.

